# Delineation of the Clinical Features and Treatment Response of Oromandibular Dystonia: A Multicenter Summary of 2,057 Cases

**DOI:** 10.1101/2020.08.28.20184085

**Authors:** Laura M. Scorr, Stewart A. Factor, Sahyli Perez Parra, Rachel Kaye, Randal C. Paniello, Scott A. Norris, Joel Perlmutter, Tobias Bäumer, Tatiana Usnich, Brian Berman, Marie Mailly, Emmanuel Roze, Marie Vidailhet, Joseph Jankovic, Mark S. LeDoux, Richard Barbano, Florence Chang, Victor Fung, Sarah Pirio Richardson, Andrew Blitzer, H.A. Jinnah, for the Dystonia Coalition Investigators

## Abstract

**Objective:** To better characterize oromandibular dystonia (OMD) to facilitate early diagnosis and test the hypothesis that botulinum toxin treatment alleviates symptoms, regardless of etiology, to provide guidance on treatment strategies.

**Methods:** To better characterize this condition we utilize a three-pronged approach. First, we provide a comprehensive summary of the world’s literature encompassing 1157 cases in 27 separate manuscripts. Next, we describe the clinical features of 727 OMD subjects enrolled by the Dystonia Coalition (DC), an international multicenter database. Finally, we provide details of the treatment approach and response from two expert centers where large numbers of OMD patients are followed. Cases from expert centers were utilized to analyze whether response to botulinum toxin varied by etiology of OMD.

**Results:** In all cohorts, typical age at onset was in the 50s and approximately 70 % of cases were female. Although the literature OMD more commonly described as a focal dystonia, analysis of the DC database revealed it more commonly appears as part of a segmental dystonia. Expert center review of 173 cases revealed botulinum toxin injections improved symptom severity by more than 50% in approximately 78% of subjects. Among the patients at expert centers, analysis revealed that treatment response did not vary by etiology.

**Conclusions:** Botulinum toxin injections are an effective treatment for OMD, regardless of etiology. By providing a more comprehensive description of OMD and the therapeutic efficacy of botulinum toxin for this type of dystonia, we hope to improve clinical recognition to aid in timely diagnosis and inform treatment strategies.

## Introduction

Dystonia is a movement disorder characterized by sustained or intermittent muscle contractions causing abnormal, often repetitive, movements, abnormal postures, or both.^1^ Oromandibular dystonia (OMD) is a relatively uncommon form that affects primarily the masticatory, lower facial, and lingual muscles, and sometimes involves other nearby muscles. The clinical presentations of OMD are varied including features of jaw opening, jaw closing, jaw deviation, abnormal lower face movements, and mixed presentations. The anatomic distribution of muscles affected varies by the OMD phenotype.^2^ It is not uncommon to observe symptoms of dystonia in adjacent cranial, cervical and pharyngeal regions given frequent segmental distribution of this form of dystonia and overlapping musculature in these anatomic regions. OMD symptoms may be task specific, triggered by speech or eating, or can be present at rest. OMD is particularly disabling because it interferes with the ability to eat and speak, and may be associated with marked discomfort.

Idiopathic focal OMD is reported to be rare, representing 3-5% of all dystonias. Incidence is estimated at 3.3/1,000,000/year and prevalence is estimated at 68.9/1,000,000.^3,4^ OMD is often unrecognized, leading to a delay in diagnosis, a delay in treatment, and sometimes inappropriate treatment. The time from onset of symptoms to diagnosis in the most common forms of dystonia can be up to 6 years, and this delay is believed to be even longer in OMD.^5,6^ Also due to its rarity, much of our understanding is based on case reports or relatively small series from expert centers. Some of these focused on idiopathic cases, others included acquired forms, and some included a high representation of inherited disorders with OMD, such as X-linked dystonia parkinsonism.^7,8,9^ The descriptions of OMD from these centers are quite varied, particularly with regards to optimal treatment strategies and outcomes. For example, some expert centers do not recommend therapy with botulinum toxin (BoNT),^10,11^ while others recommend specific strategies for its use.^7,12^ Some suggest that response of tardive and idiopathic forms of OMD to botulinum toxin is similar, but large studies are lacking.^13^ A clearer description of a large series of cases collected across multiple centers is needed to improve clinical recognition, reduce times to diagnosis, and delineate optimal treatment strategies.

The purpose of this study is to better describe the clinical characteristics and treatment response of OMD using a three-pronged approach. First, we provide a comprehensive summary of the world’s literature of this disorder, encompassing 1157 total cases in 27 separate reports. Second, we describe the clinical features of this disorder from the Dystonia Coalition (DC), a methodical international multicenter study of all types of dystonia, 727 of whom had OMD. Third, we provide details of treatments and responses of 173 cases from two expert centers and investigate whether treatment response varies by etiology. By providing a more comprehensive description of OMD and it’s response to treatment, we hope to improve clinical recognition and diagnosis of its many varied features, and comment on an appropriate treatment approach.

## Materials and Methods

### Literature Review

The PubMed database was queried from January 1989 through March 2020 for reports using the keywords ‘oromandibular dystonia,’ ‘jaw dystonia,’ ‘Meige syndrome’ and ‘lingual dystonia.’ Other reports were found through the bibliographies of these articles. Only those papers published in the English language were included. Case reports of fewer than 5 subjects were excluded. Series that described dystonia due to peripheral injury or of lower facial twisting phenomenology typical of psychogenic or functional dystonia were excluded due to diagnostic uncertainty. However, other secondary forms of OMD (e.g. tardive syndromes) were included because the majority of prior reports described combined etiologies and it was often difficult to distinguish tardive from idiopathic cases. Likewise, cases with task specific musician’s dystonia were not excluded because they could not typically be distinguished from idiopathic cases in the literature. Cases of bruxism are sometimes reported to be a feature of OMD, but the mixed pathophysiology is often inconsistent with dystonia and includes mechanical alignment issues, dental problems, disturbed sleep physiology and peripheral nervous system pathology.^14^ Therefore, such patients were only included if concurrently diagnosed with OMD. Data extracted from published case series included patient demographics and dystonia etiology, clinical features, and treatment response.

### Dystonia Coalition (DC) Cohort

Data from the DC natural history of dystonia study database were collected and analyzed for 727 OMD subjects enrolled across 26 international sites from 2011-2019 (http://clinicaltrials.gov/show/NCT01373424). To be included in the DC database the patients had to be diagnosed with isolated dystonia of any type (focal, segmental, multifocal, generalized); and inclusion for the analysis of OMD required dystonia to be present in jaw, tongue, and/or perioral region. Exclusion criteria for database entry were any evidence of a secondary cause for dystonia, significant medical or neurologic conditions that preclude completing the neurologic examination, significant physical or other condition that would confound diagnosis or evaluation, other than dystonia or tremor. Of note, presence of dystonia in contiguous body regions such as the upper face or neck was considered as a segmental pattern.

Subjects completed a 26-item questionnaire describing demographics as well as the clinical characteristics of their dystonia. A neurologist specializing in movement disorders evaluated each participant to determine distribution of dystonia, areas affected, and severity as measured on the Global Dystonia Rating Scale (GDRS).^15^ The GDRS is a Likert-like scale with which dystonia is rated from 0, if absent, to 10, if severity is maximal. Subjects were also queried about any sensory trick (also known as geste antagoniste or alleviating manoever)^16^ and prior treatments utilized. Clinical features of interest included distribution of dystonia (focal, segmental, general), areas affected (jaw, tongue, lower face), and treatments utilized. Some subjects also completed the SF 36-Item Health Survey assessing quality of life (QOL), Beck Depression Inventory II scale (BDI), and Liebowitz Social Anxiety scale (LSA). These data were collected only for those subjects with recent onset within 5 years to limit recall bias.

### Expert Center Cohorts

Data were collected from two clinical centers with expertise in the management of OMD. Retrospective chart review identified 116 additional subjects evaluated at the Emory University Movement Disorders Clinic between 2015 to 2019, and 57 subjects evaluated in Head and Neck Surgical Group (HNSG) and the NY Center for Voice and Swallowing Disorders. The primary inclusion criterion was diagnosis of OMD of any type (focal, segmental, multifocal, generalized). Secondary forms of dystonia (e.g. tardive dystonia and post-stroke dystonia) were included. Exclusion criteria were loss to follow up after initial evaluation and significant medical or physical conditions that would confound diagnosis or treatment outcomes. Demographic and clinical features including age at onset, gender, race, distribution of dystonia, and areas affected were extracted from retrospective chart review. Severity, QOL, depression, and social anxiety scores were not available for these subjects. However, additional information was collected on BoNT treatment.

### Statistical Analysis

Analyses were completed separately for each of the four cohorts (Literature review, DC, EMDC, and HNSG due to the different types of data that were available for each cohort. For each cohort, descriptive analyses for all demographics (age, sex, race) and clinical characteristics (distribution, areas affected, and severity) were completed. For the expert center cohorts with detailed treatment data, additional descriptive analysis was performed for therapies employed and response to treatment. To evaluate whether treatment response varied by etiology, ANOVA was utilized. Within the DC cohort, additional descriptive analyses were performed for BDI, LSA, and SF-36 scores for the subset of subjects within five years of onset. Univariate linear regression was performed to estimate the association between QOL and demographic/clinical characteristics. A multivariate linear regression model was constructed controlling for age and gender, as has been done in published literature assessing QOL in patients with neurologic disease. Distribution of dystonia was also controlled for as a marker of severity. All data analysis was performed with SAS version 9.4.

The Emory University Institutional Review Board, the Icahn School of Medicine, and the DC steering committee approved all procedures involving human subjects.

## Results

### Review of Published Cases

A comprehensive literature query returned 27 papers meeting our inclusion criteria. These papers describe 1,157 cases. Table 1 summarizes these reports which included idiopathic and secondary OMD. The reports were generally of single center cohorts for the purpose of defining clinical features and treatment response. Only six of the papers described cohorts larger than 50 subjects. Among these series, 68 % of subjects were female and the mean age of onset was 52. Etiology of cases varied by report, but primarily included idiopathic and tardive cases. Although it is traditional to group OMD into specific subtypes (jaw opening, jaw closing, tongue, lower face), descriptions of clinical characteristics varied considerably, with some authors defining each case by the predominant feature and many describing a mix of characteristics. The majority of reports note that subjects received a trial of BoNT, though some centers did not report treatment data or outcome or treated some subjects exclusively with oral pharmacotherapy due to perceived inefficacy of toxin injections for OMD. However, the majority of reports of subjects treated with BoNT note return for subsequent injection and subjective improvement in symptoms and/or QOL.

**Table 1.** Clinical Characteristics of 1,157 published cases of oromandibular dystonia.

| **Author** | **n** | **Etiology** | **Clinical Characteristics\*** | **BoNT trial (n)** | **Percent Improved** |
| --- | --- | --- | --- | --- | --- |
| Blitzer et al. (1989)<sup>21</sup> | 20 | NR<sup>+</sup> | NR | Yes (20) | 95% |
| Jankovic et al. (1990)<sup>22</sup> | 62 | NR | JC (NR), JO (NR) | Yes (62) | 73% |
| Hermanowicz and Truong (1991)<sup>23</sup> | 5 | NR | JO (20%), JC (80%), L (20%) | Yes (5) | 100% |
| Van den Bergh et al. (1995)<sup>24</sup> | 5 | NR | JO (80%), L (20%) | Yes (5) | 100% |
| Sankhla et al. (1998)<sup>25</sup> | 21 | NR | NR | Yes (21) | 68% |
| Tan & Jankovic (1999)<sup>26</sup> | 162 | Idiopathic (63%),<br>Tardive (22.8%),<br>Other (14.2%) | JO (21.6%), JC (52.5%), M (24%) | Yes (162) | 68% |
| Tan & Jankovic (2000)<sup>13</sup> | 116 | Idiopathic (79%)<br>Tardive (21%) | JO (23%), JC (49%), M (28%), L (20%) | Yes (116) | 100% |
| Bhattacharyya et al. (2001)<sup>27</sup> | 5 | NR | NR | Yes (5) | 100% |
| Adler et al. (2002)<sup>28</sup> | 5 | Idiopathic (100%) | JO (40%), JC (40%), M (20%) | Yes (5) | NR |
| Lo et al. (2005)<sup>29</sup> | 5 | Post-stroke (100%) | JC (100%) | Yes (5) | 33% |
| Wan et al. (2005)<sup>30</sup> | 12 | NR | NR | Yes (12) | 88% |
| Singer and Papapetropoulos (2006)<sup>31</sup> | 23 | NR | JO (52%), JC (48%) | Yes (23) | 65% |
| Lee (2007)<sup>32</sup> | 6 | NR | JC (50%), L (50%), P (67%) | Yes (2) | 100% |
| Rosales et al. (2010)<sup>7</sup> | 49 | X-linked dystonia-parkinsonism | JO (65%), JC (24%), M (12%) | Yes (49) | 100% |
| Merz et al. (2010)<sup>33</sup> | 30 | Idiopathic (83%)<br>Other (17%) | JO (10%), JC (23%), M (33%), L (10%) | Yes (30) | 100% |
| Esper, et al. (2010)<sup>9</sup> | 17 | Idiopathic (47%)<br>Tardive (41%)<br>Degenerative (6%)<br>Post-infectious (6%) | JO (53%), JC (18%), M (18%), L (100%) | Yes (9) | 78% |
| Teive et al. (2012)<sup>34</sup> | 5 | Wilson's disease (100%) | JO (100%) | Yes (5) | 100% |
| Sinclair et al. (2013)<sup>35</sup> | 59 | Idiopathic (91%)<br>Tardive (9%) | JO (36%), JC (48%) M (16%), L (17%) | Yes (59) | 100% |
| Bakke et al. (2013)<sup>36</sup> | 21 | NR | JO (19%), JC 5%), M (43%) | N R | NR |
| Termsarasab et al. (2014)<sup>10</sup> | 41 | Idiopathic (100%) | M (49%), L (12%), P (5%) | Yes (18) | 12% |
| Moscovich et al. (2015)<sup>37</sup> | 8 | NR | M (100%) | Yes (8) | 100% |
| Teemul et al. (2016)<sup>38</sup> | 6 | Idiopathic (83%)<br>Tardive (17%) | M (100%) | Yes (6) | 83% |
| Nastasi et al. (2016)<sup>39</sup> | 30 | Idiopathic (73%)<br>Tardive (13%) | JO (53%), JC (17%), M (7%), L (100%), P (27%) | Yes (30) | 87% |
| Kreisler et al. (2016)<sup>40</sup> | 14 | NR | JO (21%), JC (36%), M (7%), P (29%) | Yes (7) | (All returned for repeat injections) |
| Slaim et al. (2018)<sup>41</sup> | 240 | Idiopathic (71%)<br>Tardive (13%)<br>Other (16%) | JO (62%), JC (20%), M (27%), L (27%) | No | NR |
| Scorr et al. (2018)<sup>12</sup> | 18 | Idiopathic (83%)<br>Tardive (17%) | JO (50%), JC (17%), M (33%), L (33%), | Yes (18) | 100% |
| Yoshida (2019)<sup>42</sup> | 172 | Idiopathic (28%)<br>Tardive (62%) | JO (22%), JC (12%), M (7%), L (100%) | Yes (172) | 78% |

\*Jaw Opening (JO), Jaw Closing (JC), Mixed (M), Lingual (L), Perioral (P)
\*Not reported (NR).

### DC Cohort

Table 2 presents a cross-sectional analysis of demographics and clinical features for subjects with idiopathic OMD enrolled in the DC database. Among the 727 cases, 70% were female and the average age at onset was 50 ± 16 years. In this cohort, 87% of subjects identified as White. The distribution of dystonia was most commonly segmental (43%), and less commonly focal (39%) or generalized (10%). Sixty one percent had involvement of the jaw, 85% had involvement of the lower face, and 17% had involvement of the tongue. GDRS severity scores averaged 2.85 ± 2 for the lower face 1.84 ± 2 for the jaw and tongue, with average total scores of 16 ± 13. Among the subjects, 32% reported having received BoNT treatment. Concurrent dystonia in other regions of the body did not increase the chance that OMD patients received botulinum toxin injection therapy (p=0.53). This is consistent with prior reports that many patients do not pursue or are not offered botulinum toxin injections. There was no significant difference of exposure to treatment by distribution of dystonia (p=0.53).

**Table 2.** Clinical characteristics and treatment response of oromandibular dystonia subjects in the Dystonia Coalition database, EMDC Cohort, and Head and Neck Surgical Group (HNSG) Cohort.

| **Characteristics** | **Dystonia Coalition**<br>N=727<br>n(%) / mean (SD) | **EMDC Cohort**<br>N=116<br>n(%) / mean (SD) | **HNSG Cohort**<br>N=57<br>n(%) / mean (SD) |
| --- | --- | --- | --- |
| **Age of Dystonia Onset** | 50 (16) | 54 (15) | 50 (14) |
| *Focal* | 53 (12) | 59 (12) | NR* |
| *Segmental* | 50 (14) | 53 (14) | NR |
| *Generalized* | 26 (20) | 29 (22) | NR |
| **Sex** |  |  |  |
| *Male* | 217 (30%) | 35 (30%) | 17 (30%) |
| *Female* | 510 (70%) | 81 (70%) | 49 (70%) |
| **Race** |  |  |  |
| *White* | 634 (87%) | 76 (66%) | 20 (36%) |
| *Black* | 57 (8%) | 27 (23%) | 2 (4%) |
| *Other* | 30 (4%) | 13 (11%) | 4 (7%) |
| *Unknown* | 6 (1%) | NR | 29 (53%) |
| **Etiology** |  |  |  |
| *Idiopathic* | 727 (100%) | 75 (65%) | 46 (81%) |
| *Tardive* | NR | 25 (22%) | 11 (19%) |
| *Degenerative* | NR | 11 (9%) | NR |
| *Stroke Induced* | NR | 3 (2%) | NR |
| *Genetic* | NR | 2 (2%) | NR |
| **Distribution** |  |  |  |
| *Focal* | 284 (39%) | 53 (46%) | 46 (82%) |
| *Segmental* | 315 (43%) | 57 (49%) | 9 (16%) |
| *Multifocal* | 46 (6%) | NR | NR |
| *Hemi-dystonia* | 5 (1%) | NR | NR |
| *Generalized* | 74 (10%) | 6 (5%) | 1 (2%) |
| **Areas Affected** |  |  |  |
| *Lower Face* | 620 (85%) | 31 (29%) | 3 (5%) |
| *Jaw* | 440 (61%) | NR | NR |
| *-opening* | NR | 40 (36%) | 35 (63%) |
| *-closing* | NR | 61 (56%) | 32 (57%) |
| *-deviation* | NR | 17 (16%) | 3 (5%) |
| *-mixed* | NR | NR | 20 (36%) |
| *Tongue* | 123 (17%) | 40 (36%) | 7 (12%) |
| **GDRS Severity** |  |  |  |
| *Total* | 16 (13) | NR | NR |
| *Lower face* | 2.85 (2) | NR | NR |
| *Jaw and tongue* | 1.84 (2) | NR | NR |
| **BoNT** |  |  |  |
| *Treated* | 234 (32%) | 115 (99%) | 57 (100%) |
| *Dose (Onabotulinum toxin A equivalent units)* | NR | 102 (72) | 73 (40) |
| *EMG usage* | NR | 101 (88%) | NR |
| *Toxin Response* |  |  |  |
| *Good (75-100%)* | NR | 48 (45%) | 48 (84%) |
| *Moderate (50-74%)* | NR | 24 (23%) | 3 (5%) |
| *Partial (25-49%)* | NR | 2 (9%) | 0 |
| *Minimal (1-24%)* | NR | 19 (18%) | 1 (2%) |
| *Not reported* | NR | 1 (1%) | 5 (8%) |

\*Not Reported (NR)

Patient reported disability was measured by the SF-36 sub-scores (Table 3), which were transformed such that a score of zero is equivalent to maximum disability and a score of 100 is equivalent to no disability. Average scores represent disability in all domains, particularly in mental health (26±20), physical role (50±43), emotional role (50±14), and vitality (53±21). Mood was evaluated with BDI, on which total scores greater than 13 are indicative of depression.^17^ The average BDI score among subjects with OMD was 9.7±7.84. Anxiety was assessed using the LSA score, on which scores greater than 30 are consistent with social anxiety.^18^ The average LSA score among subjects with OMD was 33±28.

**Table 3.** Depression, Anxiety, and QOL in Oromandibular Dystonia subjects enrolled in the Dystonia Coalition Natural History Study.

| **Characteristics** | **Dystonia Coalition Natural History Study**<br>N=368<br>Mean (SD) |
| --- | --- |
| **SF-36 Sub-scores** |  |
| *Physical Function* | 71 (27) |
| *Role Physical* | 50 (43) |
| *Body Pain* | 63 (27) |
| *General Health* | 62 (22) |
| *Vitality* | 53 (21) |
| *Social Function* | 66 (29) |
| *Role Emotional* | 50 (14) |
| *Mental Health* | 26 (20) |
| **LSA Score** | 33 (28) |
| **BDI Score** | 9.7 (7.84) |

Univariate linear regression revealed age, gender, distribution and severity of dystonia as measured by GDRS were not significantly associated with total QOL score. Subjects identifying as Black had QOL scores that were worse than subjects identifying as White, indicating significantly worse physical QOL (β=-10.25, p=0.02). Mental QOL improved 0.34 points for each year older a patient was, indicating worse mental QOL for younger subjects with OMD (β=0.34, p<0.01). GDRS severity score was associated with worsened mental QOL, and each point higher on the GDRS score was associated with a 0.6-point lower mental component QOL score (β=-0.60, p<0.01). Each point higher on the BDI scale for depression was associated with a 1-point lower mental and physical component QOL score (p<0.01). Each point higher on the LSA scale for anxiety was associated with a 0.28 and 0.14-point lower score for mental and physical component QOL score, respectively (p<0.01). Reported exposure to BoNT therapy was not associated with QOL scores. These findings underscore the negative effect of OMD on QOL as well as the significance of comorbid depression and social anxiety in this population.

### Expert Center Cohorts

#### Emory Movement Disorders Clinic (EMDC)

A retrospective analysis was performed for the cohort of 116 OMD subjects evaluated and treated at EMDC within the last 5 years, who were not already enrolled in the DC (Table 2). Among these cases, 68% were female. The majority of subjects diagnosed with OMD had segmental (49%) and focal dystonia (46%) distributions, as compared to generalized (5%). The most common area affected was the jaw (63%), though involvement of the lower face (29%) and tongue (36%) were also noted. The most common movement of the jaw was closing (56%), followed by jaw opening (36%), and jaw deviation (17%). Of the subjects seen at least twice, 99% had BoNT injections. The average BoNT A equivalent unit dosage was 102±72u. EMG was used in 88% of cases to confirm injection placement. Routine clinical practice at EMDC is to record patient reported percent improvement, with 0 being no improvement and 100% being complete relief of symptoms. Subjects reported subjective improvement ranging from 50-100% in 68% of cases.

Among all cases, nine were noted to have remission (Table 4), all were women. Three were White, three Black, one Asian and two of unknown race. The mean age at onset was 54.6 ± 12.0 years, and diagnosis was 31.7 ± 35.0 months after onset. On average, 13.4 ± 9.7 sessions of BoNT therapy were performed prior to remission. The mean dose of toxin used in the last visit prior to study entry was 92.2 ± 86.1 units. The mean duration of symptoms was 6.9 ± 5.4 years and the mean duration of remission was 20.4 ± 17.3 months at the time of chart review. Seven subjects had idiopathic OMD and two were diagnosed with tardive syndrome. Eight subjects had jaw-opening dystonia, one patient had jaw deviation and one case presented a mixed form (jaw opening in addition to jaw deviation). Lingual dystonia was present in six subjects. Three participants had blepharospasm and two cervical dystonia in addition to OMD.

**Table 4.** Characteristics of remission in oromandibular dystonia cases.

| **Characteristics** | **EMDC Remission Cases**<br>N=9<br>N (%) / mean (SD) |
| --- | --- |
| **Age of Dystonia Onset** | 54.6 (12) |
| **Sex** |  |
| *Female* | 9 (100%) |
| **Race** |  |
| *White* | 3 (33%) |
| *Black* | 3 (33%) |
| *Asian* | 1 (11%) |
| *Unknown* | 2 (22%) |
| **Etiology** |  |
| *Idiopathic* | 7 (78%) |
| *Tardive* | 2 (22%) |
| **Distribution** |  |
| *Focal* | 5 (55.6%) |
| *Segmental* | 4 (44%) |
| **Areas Affected** |  |
| *Jaw* |  |
| *-opening* | 8 (89%) |
| *-closing* | 0 |
| *-deviation* | 1 (11%) |
| *-mixed* | 1 (11%) |
| *Tongue* | 6 (66.7%) |
| **BoNT** |  |
| *Treated* | 9 (100%) |
| *Dose (Onabotulinum toxin A equivalent units)\** | 92 (86) |
| **Duration of symptoms (years)** | 7 (5) |

\*Dose conversion ratio of AbobotulinumtoxinA to OnabotulinumtoxinA was 2.5:1.

#### Head and Neck Surgical Group Cohort (HNSG)

A retrospective analysis was performed on 57 OMD subjects evaluated at HNSG within the last five years (Table 2). Among these cases, 70% were female. The majority had focal dystonia (82%), as compared to segmental (15%) and generalized (2%). The most common area affected was the jaw and presentations included jaw opening (63%), jaw closing (57%), jaw deviation (5%), and mixed phenomenology (36%). Lingual dystonia was not reported. All subjects received BoNT and the average onabotulinumtoxinA equivalent dosage was 73 ± 40u. Patients reported improvement in pain and/or function as good, per physician notes, in 84% of cases.

#### Evaluation of Differences in Clinical Features and Treatment Response by Etiology

To evaluate whether clinical characteristics and BoNT treatment response varied by etiology, the data from EMDC and the HNSG were pooled for analysis. Etiology of OMD was categorized as Idiopathic, Tardive, or Other (degenerative, post-stroke, related to a genetic syndrome such as Wilson’s disease or PKAN). Clinical characteristics and treatment responses were investigated to determine whether it was a function of etiology (Table 5). Demographic features including age of onset and gender distribution did not vary significantly by etiology. Features of lingual dystonia were more common among tardive cases (p=0.03), but there was no significant difference in the occurrence of other phenomenologies by etiology. Also, total toxin dose (p=0.78) and toxin response did not vary by etiology (p=0.56).

**Table 5.** Variation in clinical characteristics and treatment response of oromandibular dystonia by etiology in expert center cases.

| **Characteristics** | **Idiopathic**<br>N=121<br>n(%) / mean (SD) | **Tardive**<br>N=36<br>n(%) / mean (SD) | **Other**<br>N=16<br>n(%) / mean (SD) | **p-value** |
| --- | --- | --- | --- | --- |
| **Age at Dystonia Onset** | 54 (13) | 54 (15) | 46 (23) | 0.97 |
| **Sex** |  |  |  | 0.54 |
| *Male* | 36 (30%) | 9 (30%) | 6 (40%) |  |
| *Female* | 85 (70%) | 27 (74%) | 10 (60%) |  |
| **Areas Affected** |  |  |  |  |
| *Lower Face* | 33 (27%) | 8 (21%) | 3 (20%) | 0.76 |
| *Jaw* |  |  |  |  |
| *-opening* | 59 (49%) | 14 (40%) | 4 (26%) | 0.15 |
| *-closing* | 64 (53%) | 21 (57%) | 13 (80%) | 0.09 |
| *-deviation* | 17 (14%) | 4 (11%) | 1 (5%) | 0.56 |
| *Tongue* | 33 (27%) | 15 (43%) | 2 (10%) | 0.03 |
| **BoNT** |  |  |  |  |
| *Dose (Onabotulinum toxin A equivalent units)* | 88 (64) | 73 (46) | 90 (75) | 0.48 |
| *Toxin Response* |  |  |  | 0.56 |
| *None* | 5 (4%) | 0 | 0 |  |
| *>50% Improvement* | 96 (79%) | 27 (76%) | 13 (80%) |  |

## Discussion

In this study, we summarize the clinical characteristics and treatment of OMD in the largest combined cohort reported, 2,057cases. This included 1,157 cases from the world’s literature, 727 cases from an international multicenter biorepository, and 173 cases treated at two expert centers. The data from these three cohorts were not uniform. The cohort from the DC included only idiopathic OMD, while cases from the literature and the 2 expert centers included secondary cases. Anatomic treatment distribution of dystonia was reported by the DC and expert centers, but not consistently in the literature. Treatment data were described most thoroughly by the expert centers. Despite these differences among the cohorts, overall results were strikingly similar. OMD tends to occur in the early 50s and is more common in women and more common in whites compared to other races.

The literature on OMD is comprised primarily of idiopathic cases, though secondary cases occurring because of tardive, genetic, degenerative, and lesion-based etiologies have been reported in smaller numbers. The approach to diagnosis and characterization of OMD is varied. Some centers focus on the predominant movement abnormality, while others describe mixed phenomenology by presence or absence of involvement of muscle groups. Assessment of OMD severity also varies from clinical impression, to patient reported outcomes such as the Oromandibular Dystonia Questionnaire (OMDQ-25), to use of general dystonia scales such as the GDRS. These varied methods make comparisons of patient populations across reports challenging and highlight the need for development of a disease specific severity scale to be used for standardized characterization of phenomenology, severity, and treatment response. Although the OMDQ-25 provides valuable patient reported outcomes, this scale is influenced by mood and may not be specific to motor symptoms. Despite the heterogeneity of etiology and phenomenology, most cases improved with BoNT treatment. Of the 23 manuscripts reporting treatment response, 11 reported 100 % of cases improved, 10 reported greater than 65 % of cases improved, and only two reported less than 50% of cases improved (Table 1). Future work is necessary to determine whether the phenomenology or injection approach plays a role in response.

There several limitations to our literature review, including relatively small subject samples from single centers. Of the 27 reports summarized in Table 1, only 4 describe cohorts greater than 100 subjects. The analysis using the DC multicenter international database minimizes the idiosyncrasies often present in single center analyses providing valuable information on the clinical characteristics of subjects diagnosed with OMD from a database of clinical characteristics of all types of dystonia. Interestingly, review of the DC database reveals that OMD was typically present as part of a segmental pattern of dystonia involving a contiguous body region such as the upper face and neck. Most published reports have described OMD primarily as a focal dystonia. This finding draws attention to the need to carefully evaluate subjects with blepharospasm and cervical dystonia for dystonia in the oromandibular region. Given our finding that OMD commonly occurs in combination with other forms of dystonia and previous reports that 50% of subjects with blepharospasm have spread of dystonia to the oromandibular region,^19^ it is possible that the oromandibular region is particularly sensitive to dystonia spread. Future longitudinal studies are needed on the natural history of OMD to assess likelihood of progression of dystonia to other body regions.

Within the DC cohort, regions most commonly affected in OMD were lower face (85%), followed by jaw (61%) and tongue (17%). Many subjects had a combination of regions involved suggesting that it will be important for future disease specific rating scales and treatment algorithms to address mixed phenomenology. Severity of dystonia was moderate in this cohort with an average total GDRS score of 16 ± 13, and severity in the lower face of 2.85 ± 2 and jaw and tongue of 1.84 ± 2. The marked effect on QOL despite relatively low average scores for regions involved in OMD, suggests OMD severity may not be adequately captured by GDRS scoring. Despite moderate severity and average duration of dystonia of 11 ± 12 years at the time of enrollment, only 32% reported treatment with BoNT. Furthermore, in this population, despite low to moderate severity as measured on GDRS, OMD seems to disproportionately affect QOL with subjects reporting poor QOL as measured by SF-36, social anxiety as measured by LSA (33 ± 28), and depression as measured by BDI (9.7 ± 7.8). Interestingly, exposure to BoNT treatment did not significantly alter QOL scores in the DC cohort. Given results that anxiety and depression significantly affect QOL, it will be important to assess for these non-motor features of disease in clinical care and to address these symptoms as part of the treatment plan for subjects. One limitation of treatment data collected in the DC database is the cross-sectional nature, without details of treatment dose or injection pattern. Given centers enrolling subjects are typically tertiary care centers, there may be an enrollment bias such that subjects presenting at these centers have failed routine treatment approaches in the community. A small prospective trial reported improved QOL after BoNT treatment at an expert center, indicating further longitudinal studies are needed.^12^

Review of expert center data from EMDC and HNSG provide more detailed information on toxin response. The demographic distribution of subjects cared for in expert centers was similar to that reported in the literature and the DC. In both centers, most subjects were treated with BoNT injections. Most subjects reported greater than 50 % improvement in symptoms. At both centers, less than 10% of subjects reported no improvement. Interestingly, the treatment response was not significantly associated with etiology with 79% of idiopathic and 76 % of tardive OMD patients reporting >50% improvement in symptoms. This finding may be a reflection of the gold standard treatment with BoNT treatment being a symptomatic treatment that may not be directly related to disease pathogenesis. A limitation of these data is that outcomes are retrospective and based on subjective patient report, which may be susceptible to bias. Routine use of a disease specific rating scale would provide more objective data. Additional prospective trials are needed to confirm optimal dose and injection pattern of BoNT for OMD.

Review of OMD cases treated within the last five years at EMDC revealed several cases of remission (Table 4). The demographic data of these cases was similar to the larger cohort, though all cases of remission occurred in women. All subjects were treated with BoNT prior to remission. Unlike with cervical dystonia ^20^ remission in OMD occurred after mean disease duration 6.9 years.

A particular strength of this study is that it includes by far the largest cohort of OMD subjects reported. Our analysis provides valuable information on the clinical characteristics of subjects diagnosed with OMD, which has been largely neglected in the literature. Additionally, the use of a multicenter international database minimizes the idiosyncrasies often present in single center analyses. This is also the first study to examine the influence of psychiatric features in subjects with OMD, further expanding our knowledge of the disease. Furthermore, review of expert center cases provided greater insight into treatment methods and prognosis. Finally, the expert center cases provided an opportunity to address whether disease etiology affects treatment outcomes. The finding that OMD responds well to BoNT regardless of etiology may encourage more widespread use of this therapy.

Future directions include investigating the natural history of OMD to determine predictors of progression or remission of symptoms. To better track dystonia severity over time, it will be necessary to develop a disease specific severity rating scale. Additionally, we must adapt the clinical assessment of these subjects to include screening for mood disturbances, such as depression and anxiety, which were found to be major determinants of QOL in this population. Finally, it is critical to clarify the best treatment strategy for OMD given the results of this analysis showing its negative impact on QOL. Prospective controlled trials are needed to confirm the efficacy of BoNT treatment and clarify best treatment methods.

## Data Availability

The data utilized from the Dystonia Coalition database is openly available by request.

## Acknowledgements

DC investigators that contributed to the recruitment of subjects for these analyses included Alan Freeman, Alberto Espay, Alex Pantelyat, Alfredo Berardelli, Allison Brashear, Andres Deik, Brian Berman, Christine Klein, Claudia Testa, Cynthia Comella, Daniel Truong, Emmanual Flamand Roze, Fatta Nahb, Francis Walker, Ihtsham Haq, Irene Malaty, Hyder A. Jinnah, Joseph Jankovic, Joel Perlmutter, Julie Leegwater-Kim, Laura Scorr, Lawrence Severt, Lydy Shih, Mahlon DeLong, Mark Hallet, Mark S. LeDoux, Natividad Stover, Oksana Suchowersky, Philipp Capetian, Pinky Agarwal, Pravin Khemani, Rachel Saunders-Pullman, Ramon Rodriguez, Richard Barbano, Sarah Richardson, Stephen Grill, Stephen Reich, Stewart Factor, Susan Fox, Sylvain Chouinard, Tanya Harlow, Tao Xei, Victor Fung, William Ondo, and Zoltan Mari.

